# Compensatory drive and sleep-related phantom limb pain: the moderating role of autonomic reserve

**DOI:** 10.64898/2026.10.06.26364856

**Authors:** Nadine Aranis, Marcela Holmes, Anna Lepesteur Gianlorenco, David Sparrow, David Crandell, Wolnei Caumo, Felipe Fregni

**Author notes:** **Correspondence to:** F. Fregni, Neuromodulation Center and Center for Clinical Research Learning, Spaulding Rehabilitation Hospital and Massachusetts General Hospital, Harvard Medical School, 96 13th Street, Charlestown, Boston, MA, USA. **E-mail address:** (F. Fregni).

## Abstract

Phantom limb pain (PLP) is common after amputation, yet why poor sleep becomes pain in only some individuals is unknown. We hypothesized that a regulatory response is interpretable only against the reserve sustaining it, and thus we tested whether autonomic capacity relative to demand moderates the sleep–pain relationship in 121 amputee adults. Sleep was assessed with the Pittsburgh Sleep Quality Index; PLP, residual limb pain (RLP), and phantom limb sensation (PLS) on 0–10 scales. Heart rate variability was summarized as autonomic reserve (log-transformed SDNN, RMSSD, total and high-frequency power) and autonomic drive (normalized low-frequency power). Poorer sleep was associated with greater PLP and RLP, but not PLS. No single HRV index moderated these associations, but the sleep × reserve × drive interaction did for PLP (b = −0.51, p = 0.026). Poor sleep predicted pain most strongly under high drive with reduced reserve. PLP vulnerability reflects autonomic capacity relative to demand, instead of any single HRV marker alone.

## Introduction

Individuals following amputation may experience several sensory and pain-related consequences, including phantom limb pain (PLP), non-painful phantom limb sensation (PLS), and residual limb pain (RLP)^1^. Although these experiences frequently coexist, they are clinically distinct. PLP is perceived in the missing portion of the limb, RLP is localized to the residual limb, and PLS refers to phantom experiences that are not painful. Particularly, PLP affects approximately 64% of people with limb amputation and represents a substantial source of disability and treatment burden^2^. Response to available interventions is frequently inconsistent, and patients are often exposed to multiple rounds of ineffective treatment before finding relief^3,4^. This variability may reflect the multifactorial nature of post-amputation phenomena, which can be influenced by clinical, psychological, behavioral, and physiological factors, mood symptoms, sleep disturbance, time since amputation, medication use, and alterations in autonomic regulation^5^. Among these factors, sleep disturbance is particularly common and may interact with physiological regulatory mechanisms to influence the intensity and persistence of PLP.^6,7^

Sleep and chronic pain have a reciprocal and potentially self-reinforcing relationship. Poor sleep can increase pain sensitivity, impair endogenous pain modulation, and predict greater pain on the following day, whereas pain can disrupt sleep initiation, continuity, and restorative quality^8,9^. Evidence suggests that sleep may be a stronger predictor of subsequent pain than pain is of subsequent sleep disturbance in many chronic pain conditions such as fibromyalgia, migraines, and arthritis populations^8^. However, the strength of this association varies considerably across individuals, suggesting physiological factors moderate the extent to which poor sleep translates into pain^7^. In the amputee population specifically, this relationship remains understudied. Although sleep disturbance has been documented among individuals with limb loss, available evidence is predominantly cross-sectional^10^, and the few longitudinal studies that have considered sleep have generally treated it as a component of quality of life or overall clinical burden instead of as a variable examined for its own temporal relationship with post-amputation pain^11,12^.

Sleep-promoting neurons are anatomically and functionally tangled with the central autonomic network, and the sleep–wake cycle is accompanied by pronounced shifts in autonomic balance. This bidirectional coupling means that abnormal sleep can produce a secondary imbalance in sympathetic drive, positioning autonomic regulation as a physiologically plausible link between sleep and pain^13,14^. Further evidence demonstrates that the autonomic nervous system (ANS) remains responsive to nociceptive input during sleep^15^. In healthy adults, nociceptive stimulation produced cardiac acceleration and increased low-frequency HRV activity across all sleep stages, with stronger responses when stimuli elicited cortical arousal^16^. These findings suggest that autonomic responses to nociception are preserved during sleep but may be amplified by sleep-dependent cortical activation.

Heart rate variability (HRV) provides a non-invasive index of cardiac autonomic regulation. Altered HRV has been reported in individuals with poor sleep^17^ and in amputees, independent of PLP status^18^. Although evidence in amputees specifically remains limited, we recently found that poorer sleep quality in individuals with PLP was associated with time-domain indices of autonomic function^19^. Prior work has, however, typically examined individual HRV indices one at a time. Time- and frequency-domain measures are highly correlated and largely redundant^20^, and analyzing them independently obscures a distinction that is central to regulatory physiology: the difference between a system with sufficient capacity to accommodate a demand and one that is mounting the same response from a depleted capacity^21^. Composite HRV indices have been used to summarize correlated measures and reduce reliance on any single parameter^22,23^ but a composite alone still yields a single number and therefore cannot represent capacity and demand as separate quantities.

The framework we adopt here follows a general logic of physiological compensation that we have recently formalized for the motor cortex in chronic pain^24^. In any regulatory system, the output of a compensatory response cannot be interpreted without reference to the capacity from which it is drawn. The canonical example is glucose regulation: the same elevated insulin concentration indicates successful β-cell compensation when glycemia is normal and β-cell failure when glycemia is high, which is why insulin resistance is quantified as insulin relative to glucose rather than as insulin alone^25,26^. The same principle underlies cognitive reserve, in which equivalent neuropathology produces markedly different clinical expression depending on the capacity available to compensate for it^27^, and allostatic load, in which a response that is adaptive when transient becomes damaging when it must be sustained beyond the capacity of the system^28^. Applied to chronic pain, we have shown that motor cortex inhibition that is analgesic in patients with preserved neurocompensatory capacity loses its efficiency in patients carrying a high burden of comorbid stressors, so that the same inhibitory signature indicates effective regulation in one patient and compensatory saturation in another, and that this efficiency is restored by successful rehabilitation^24^. The link between HRV and compensatory capacity is, moreover, not merely metaphorical: lower HRV predicts poorer β-cell compensation for insulin resistance in vivo^22^, suggesting that autonomic variability indexes a general regulatory reserve rather than a cardiac property alone.

Building on this framework, we conceptualized autonomic state along two complementary dimensions: reserve and drive. Autonomic reserve denotes the overall magnitude of variability available to accommodate physiological demand, reflecting the system’s functional capacity. Autonomic drive denotes the relative predominance of low-frequency organization within the autonomic response, interpreted within this framework as an index of ongoing physiological demand. By analogy with metabolic regulation, drive corresponds to insulin secretion, whereas reserve corresponds to the β-cell capacity supporting that secretion. Elevated drive in the context of preserved reserve is thus conceptualized as effective compensation, whereas comparable drive in the context of diminished reserve represents compensatory strain, analogous to the increased secretory demand associated with insulin resistance. The physiological significance of each dimension is therefore conditional on the other, making their interaction central to evaluating the framework. This formulation yields a directional prediction beyond a general association between reduced heart rate variability and adverse outcomes: diminished reserve is expected to strengthen the association between sleep disturbance and pain primarily when autonomic drive is elevated. Under lower drive, physiological demand may be insufficient to reveal limitations in reserve, and its association with the sleep–pain relationship is consequently expected to be attenuated or absent.

We therefore tested whether autonomic function, indexed by HRV, moderates the relationship between sleep quality and post-amputation pain in adults with limb amputation. The primary outcome was PLP, with RLP and PLS examined as secondary outcomes to assess specificity. We hypothesized that the association between sleep quality and PLP would depend on the interaction between autonomic reserve and drive, rather than on either dimension considered independently, and that the sleep–pain association would be steepest when reserve was reduced and drive elevated.

## Methods

### Study design and participants

This cross-sectional secondary analysis used baseline, pre-intervention data from an ongoing parallel-group randomized clinical trial titled “Pragmatic Trial of Remote tDCS and Somatosensory Training for Phantom Limb Pain with Machine Learning to Predict Treatment Response” (PLP-EVEREST), being conducted in the Neuromodulation Center, Spaulding Rehabilitation Hospital (Boston, MA, USA), following the local research ethics regulations and the Declaration of Helsinki. The parent trial was approved by the Mass General Brigham Institutional Review Board (protocol 2021P003323) and is registered at ClinicalTrials.gov (NCT 05188183). We enrolled adults from across the United States if they were 18 years of age or older, had an acquired amputation of one or more limbs, had current chronic pain with an average pain intensity of at least four on the visual analogue scale (VAS, 0: no pain, 10: worst imaginable pain), and experienced PLP regularly (at least once a week). Participants were required to sign the informed consent form to be included in the study. Subjects were excluded if they had any clinically significant, unstable medical or psychiatric disorder, history of substance abuse in the past 6 months (according to the DSM-5 criteria for substance use disorder with six or more symptoms), uncompensated psychiatric disorder, previous significant neurological history with current significant neurological deficits, or previous neurosurgical procedure with craniectomy.

### Exposure, outcome and covariates

Sleep quality was assessed using the Pittsburgh Sleep Quality Index (PSQI), a 19-item questionnaire producing a global score ranging from 0 to 21, with higher scores indicating poorer sleep quality^29^. Three post-amputation symptom outcomes were recorded on 0–10 visual analogue scales (VAS): PLP, RLP, and PLS. PLP was designated the primary outcome. PLS was included as a non-painful comparison outcome because it arises from the same deafferented territory but lacks the aversive dimension. Its inclusion allowed us to explore whether any observed moderation was specific to pain or extended to phantom experiences more broadly. Depressive symptoms measured by the Beck Depression Inventory (BDI), anxiety measured by the Beck Anxiety Inventory (BAI), use of current pain medication (no/yes), and time since amputation in years were recorded and used in sensitivity analyses.

### HRV acquisition and processing

RR intervals (RRi) were recorded using a Polar H10 band positioned on the patient’s chest, at the lower third of the sternum. A trained physician oversaw the procedure, ensuring participants remained at rest for at least 5 min and breathed at their normal rate.

RRi data were exported to Microsoft Excel for initial quality control. The mean RRi was calculated from the central portion of each recording, and intervals below 80% or above 120% of this mean were identified as potential artifacts or ectopic beats. Flagged intervals were removed and replaced with the mean of the preceding and following acceptable intervals. Recordings requiring correction of more than 20% of RR intervals were excluded. A stable 300-second segment was then selected for HRV analysis, following the Task Force of the European Society of Cardiology and the North American Society of Pacing and Electrophysiology recommendations^30^. The resulting RRi series were analyzed using Kubios HRV software (version 2.2; Kuopio, Finland), where frequency-domain parameters were estimated using fast Fourier transform spectral analysis with a 256-second window. Time-domain indices included: (1) standard deviation of normal-to-normal RRi (SDNN), (2) root mean square of successive RRi differences (RMSSD), and (3) percentage of successive normal RR intervals more than 50 ms (pNN50). Frequency-domain measures included total power (TP), low-frequency power (LF), and high-frequency power (HF), expressed in absolute units (ms^2^), as well as normalized LF power (LFnu).

Candidate measures for the reserve index were restricted to HRV-linear parameters. SD1 and SD2 were not included because they are redundant with RMSSD and SDNN, respectively; pNN50 was excluded because of substantial floor effects; and DFA α_1_ was excluded because it reflects fractal organization rather than HRV magnitude.

### Conceptual framework

The analysis operationalizes the compensation model described in the Introduction, in which the extent to which a physiological perturbation translates into symptom expression is governed by two distinct properties of the regulatory system: the reserve available to buffer the perturbation and the drive currently being mounted against it. Under this model, neither quantity is expected to be fully informative alone. A large compensatory response may be adaptive when reserve is sufficient to sustain it but indicates decompensation when reserve is limited, in the same way that an insulin concentration is interpretable only against concurrent glycemia. Accordingly, reserve and drive were modeled jointly, with their three-way interaction with sleep quality representing the single primary test of the model; the constituent main effects and two-way interactions were retained to preserve model hierarchy. Because reserve and drive contain related frequency-domain information, their interaction could partly reflect mathematical coupling rather than physiological interaction. We therefore quantified the correlation between the indices and repeated the primary analysis using alternative reserve definitions that excluded overlapping frequency-domain components.

### Autonomic reserve

Reserve was defined as global HRV magnitude and calculated from four time- and frequency-domain indices: SDNN, RMSSD, TP, and HF power. Because these measures were strongly right-skewed, each was natural-log transformed and standardized before being averaged:

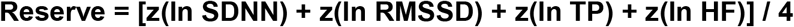

This was analyzed continuously in all primary inferential models. Higher scores indicate greater autonomic reserve, reducing dependence on any single measure. The diagnostic statistics supporting its construction are reported in Supplementary Table 1. Absolute LF power was not included in the reserve index because it forms the numerator of the drive index described below; its inclusion would therefore have caused the two axes to share a term directly. HF was retained because vagally mediated variability is conceptually part of autonomic reserve, although it also contributes to the denominator of LFnu. Consequently, reserve and drive were not completely independent (r = −0.17; Supplementary Figure 1). The primary interaction was repeated using prespecified alternative reserve definitions, including an index without HF, each reserve component individually, and a reserve score residualized with respect to the drive (Supplementary Table 2).

### Autonomic drive

Drive was indexed by LF power expressed in normalized units (LFnu):

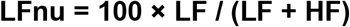

LFnu was selected as the primary drive index because it is bounded and was approximately symmetric in this sample (skewness −0.75; range, 23.0–97.2). Because LFnu is a monotonic transformation of the LF/HF ratio, the two measures contain essentially the same relative spectral information and were not included simultaneously. As a sensitivity analysis, the primary three-way interaction model was repeated after replacing LFnu with ln(LF/HF) to evaluate whether the findings depended on the scale used to represent drive. Conclusions were unchanged.

## Statistical analysis

Participant characteristics, sleep quality, post-amputation symptom outcomes, and HRV measures were summarized using means and standard deviations or medians and interquartile ranges, as appropriate. Categorical variables were summarized using frequencies and percentages. Sample size was determined by enrolment in the parent trial; no a priori power calculation was performed for this secondary analysis. Analyses were performed using available complete cases. Separate ordinary least-squares regression models were fitted for PLP, RLP, and PLS. PSQI score was the main exposure, and initial models were adjusted for age and sex. Fully adjusted models additionally included depressive symptoms, anxiety symptoms, current pain-medication use, and time since amputation. Pearson correlations were used to examine the relationships of PSQI with BDI and BAI. Because RLP exhibited a substantial concentration of zero values, PSQI scores were compared between participants with and without RLP using a two-sample t-test. Among participants reporting RLP, the association between PSQI and RLP intensity was assessed using Spearman’s rank correlation.

Moderation models additionally adjusted for resting heart rate because all HRV magnitude indices were associated with heart rate (r = −0.33 to −0.40). Without this adjustment, the reserve index could partly reflect variation in heart rate rather than autonomic variability. Continuous predictors included in these models were standardized as z-scores, while symptom outcomes remained on their original 0–10 VAS scales. Moderation was tested in two stages. First, each HRV index was entered individually as a moderator of the sleep–post-amputation symptom slope, giving 15 interaction tests (five indices × three outcomes). These models assessed whether any individual HRV measure moderated the association independently. Second, reserve and drive were entered jointly in a three-way interaction with PSQI, with all constituent main effects and two-way interaction terms (PSQI × reserve, PSQI × drive, and reserve × drive). The three-way interaction was the principal test of moderation.

For interpretation, conditional PSQI slopes were converted back to VAS points per one-point increase in PSQI. The three-way interaction was decomposed by estimating differences in the PSQI–post-amputation symptom slope across levels of reserve and drive. Reserve and drive were dichotomized at their respective sample medians only for visualization and descriptive subgroup analyses. These median-defined analyses did not replace the primary continuous interaction test, and within-subgroup coefficients and p-values were interpreted descriptively. Sensitivity analyses examined the stability of the PLP three-way interaction after sequential adjustment for depressive symptoms, anxiety, current pain-medication use, and time since amputation. The interaction was also evaluated using individual reserve components and alternative composite definitions of reserve, after replacing LFnu with ln(LF/HF) as the drive index and using a percentile bootstrap with 3,000 resamples.

PLP was the prespecified primary outcome and the PSQI × reserve × drive interaction the single prespecified primary test; RLP and PLS were secondary outcomes included to assess specificity, and the single-index and median-stratified analyses were descriptive. Because inference rests on one hypothesis-driven test whose robustness was examined through sensitivity analyses rather than on a family of independent comparisons, nominal two-sided p-values are reported throughout without adjustment for multiplicity. Statistical significance was defined as p < .05. Analyses were conducted using R Statistical Software, version 4.6.1.

## Results

### Sample characteristics

Complete sleep and HRV data were available for 121 of 122 participants; one was excluded for a missing PSQI. Mean age was 57.6 years ± 15.43, and 42 participants (34%) were female. Sleep quality was poor overall: mean PSQI 8.28 ± 4.24, with 80 participants (66%) scoring above the conventional cut-off of 5. Mean post-amputation symptom intensities were 5.43 ± 2.54 for PLP, 3.09 ± 3.11 for RLP, and 4.68 ± 3.38 for PLS. Median SDNN was 18.4 ms, and median LFnu was 74.02 (Supplementary Table 3). Participants above and below the median of the reserve index differed markedly on every constituent measure (median SDNN 31.2 vs 10.9 ms; RMSSD 24.3 vs 8.8 ms; TP 814 vs 105 ms^2^; HF power 202 vs 20 ms^2^, Supplementary Table 4).

### Poorer sleep is associated with phantom pain but not with phantom sensation

Higher PSQI scores were associated with greater PLP intensity (b = +0.155 per PSQI point, p-value = 0.004) and greater RLP (b = +0.177, p-value = 0.008), with no association with PLS intensity (b = +0.079, p-value = 0.273; Table 1, Figure 1). Both pain associations were essentially unchanged after adjusting for depressive symptoms, anxiety, use of current pain medication, and time since amputation, whereas the phantom sensation association attenuated further. PSQI correlated only weakly with BDI (r = 0.15) and BAI (r = 0.21), indicating that the sleep–pain relationship is not accounted for by affective symptom burden.

**Table 1.**
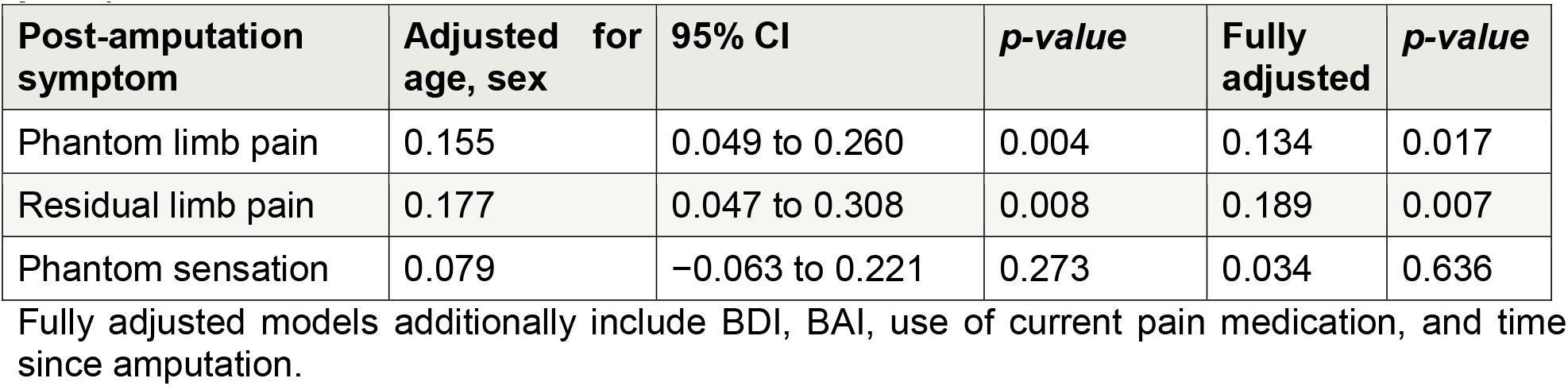
Association between sleep quality and each symptom (VAS points per PSQI point).

| Post-amputation symptom | Adjusted for age, sex | 95% CI | *p*-value | Fully adjusted | *p*-value |
| --- | --- | --- | --- | --- | --- |
| Phantom limb pain | 0.155 | 0.049 to 0.260 | 0.004 | 0.134 | 0.017 |
| Residual limb pain | 0.177 | 0.047 to 0.308 | 0.008 | 0.189 | 0.007 |
| Phantom sensation | 0.079 | −0.063 to 0.221 | 0.273 | 0.034 | 0.636 |

Fully adjusted models additionally include BDI, BAI, use of current pain medication, and time since amputation.

**Figure 1.**
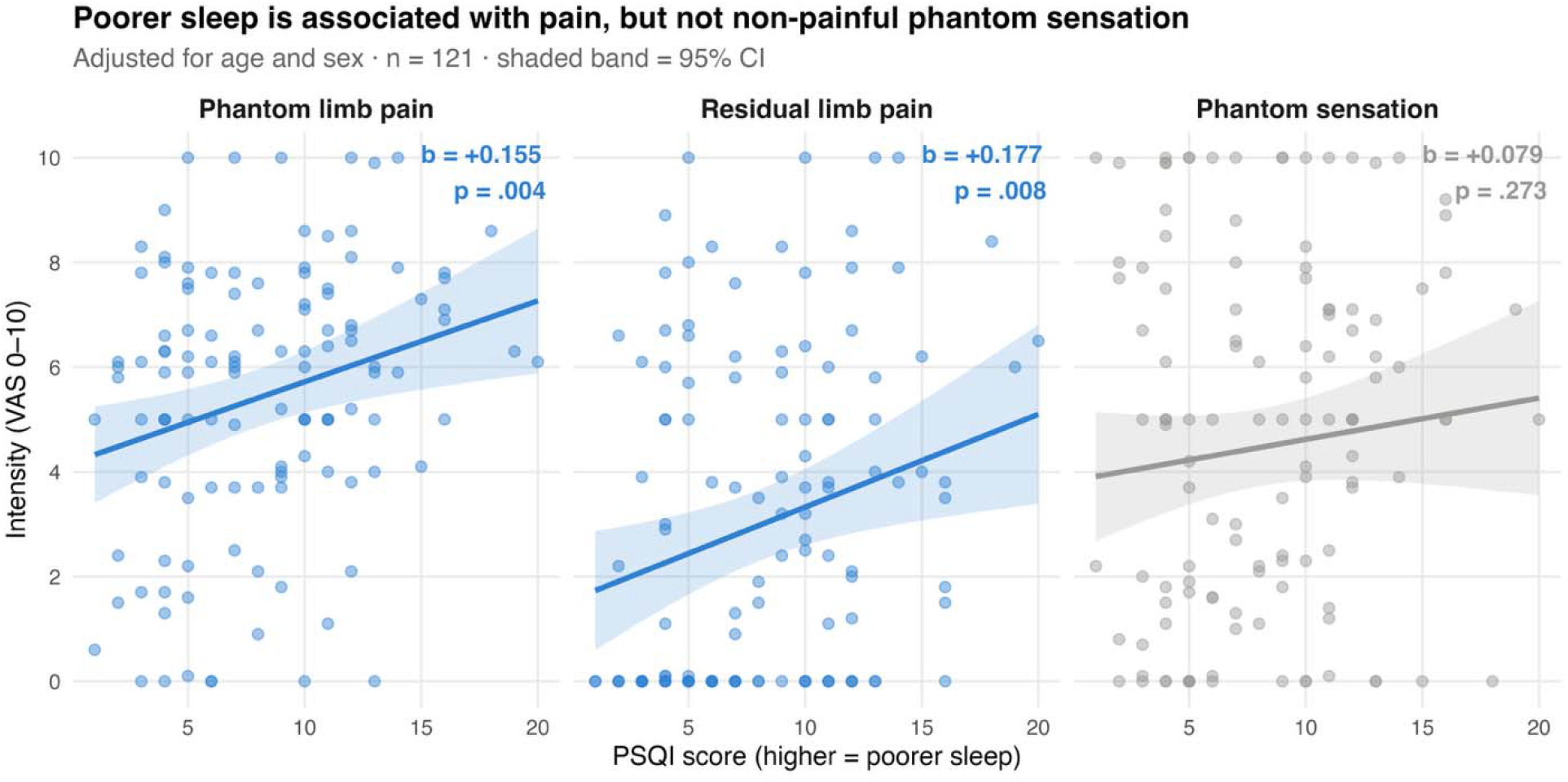
Sleep quality and post-amputation symptoms. Figure 1: Points represent observed values. Lines represent predicted values from models adjusted for age and sex, and shaded bands indicate 95% CI. The residual limb pain panel shows a concentration of observations at zero. PSQI, Pittsburgh Sleep Quality Index; VAS, visual analog scale.

### The residual limb pain association reflects occurrence rather than severity

Residual limb pain was absent in 43 of 121 participants (36%). Decomposing the association showed that it was carried entirely by whether RLP pain was present rather than by how intense it was. Participants reporting any RLP had substantially poorer sleep than those reporting none (PSQI 9.15 ± 4.26 vs 6.70 ± 3.76; t = 3.16, p-value = 0.002), whereas among affected participants PSQI was unrelated to intensity (Spearman ρ = 0.05, p-value = 0.67; Figure 2). The linear coefficient in Table 1 should therefore be read as a threshold, and not as a dose– response effect.

**Figure 2.**
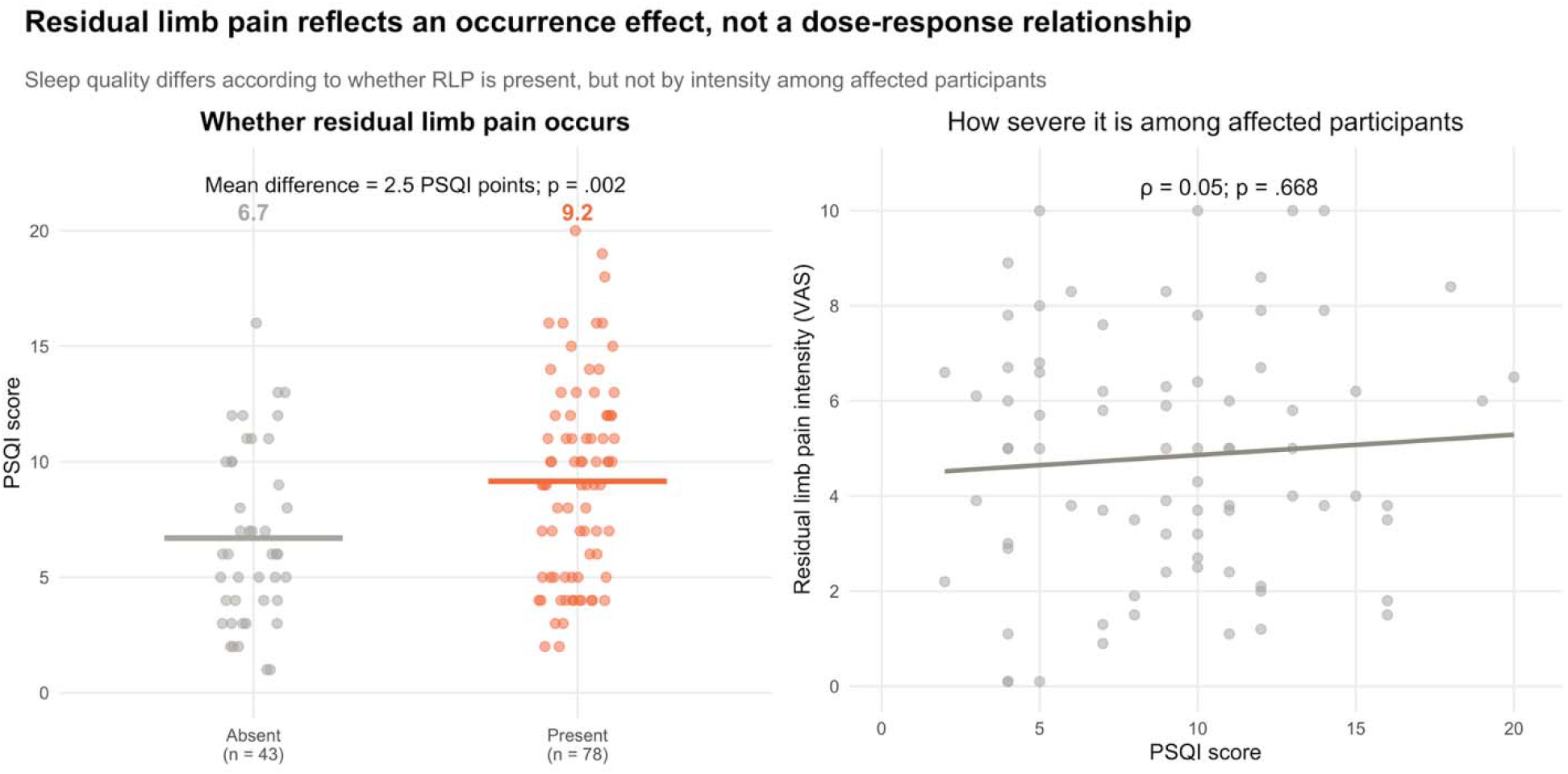
Sleep quality according to residual limb pain occurrence and severity. Figure 2: The left panel presents PSQI scores according to whether RLP was absent or present; horizontal lines indicate group means. The right panel presents the association between PSQI and RLP intensity among participants reporting any RLP. The fitted line is included for visualization, whereas ρ (rho) and its p-value are based on Spearman correlation. *PSQI, Pittsburgh Sleep Quality Index; RLP, residual limb pain; VAS, visual analog scale*.

### No single autonomic index moderates the sleep–pain relationship

Each HRV index was first entered individually as a moderator of the association between sleep and post-amputation symptom, producing 15 interaction tests. None of the individual interaction terms were statistically significant, and all 95% CIs included zero; the smallest p-value across all was 0.26 (Figure 3). The same pattern was observed when the HRV indices were analyzed on their untransformed scales, indicating that the null findings were not attributable to the right-skewed distributions of the raw HRV measures. In contrast, the joint reserve × drive interaction excluded zero for PLP.

**Figure 3.**
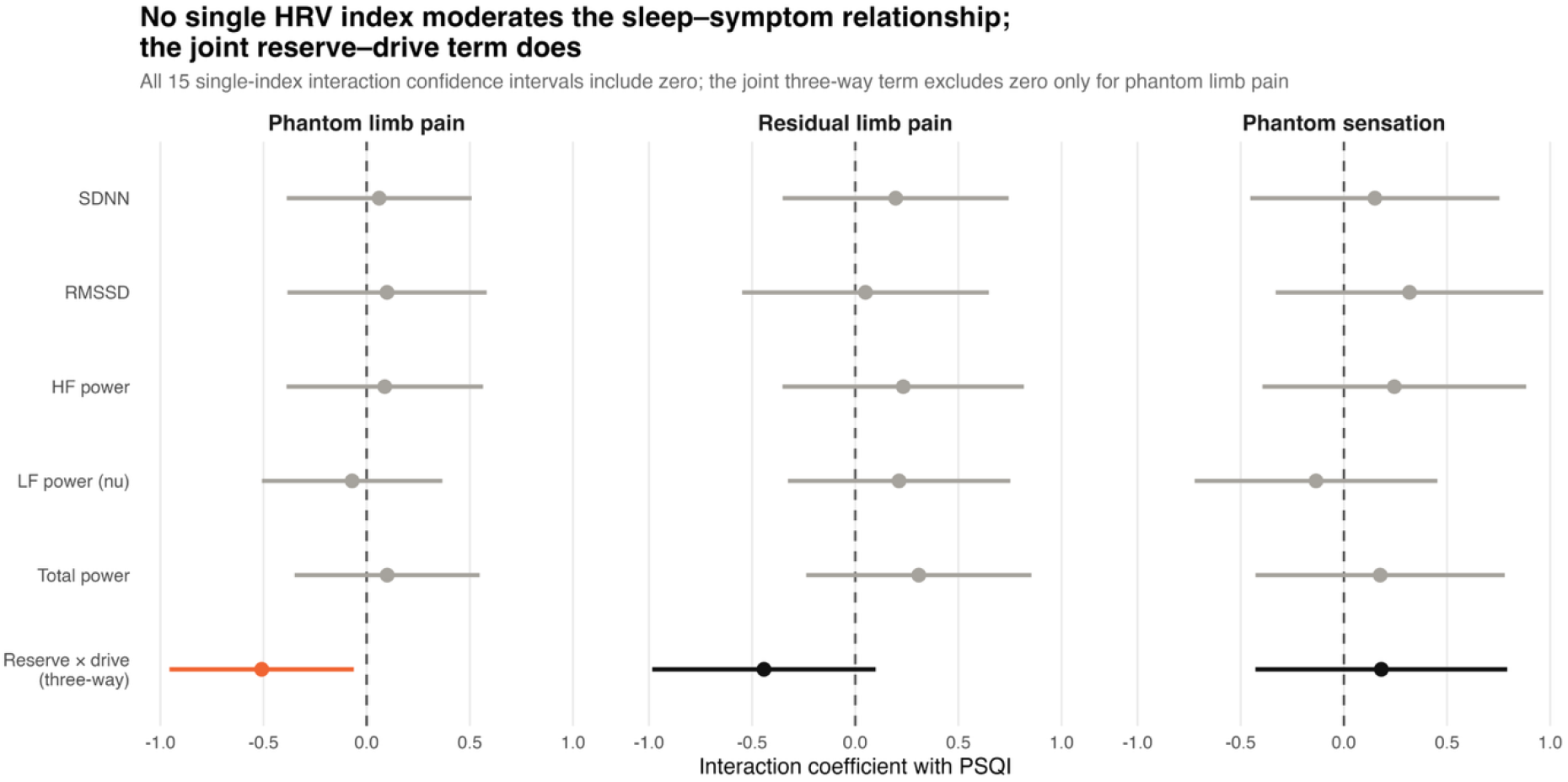
Interaction estimates with PSQI for each single HRV index and for the joint reserve × drive term, by outcome. Figure 3: Points represent interaction coefficients, and horizontal lines represent 95% CIs. The reserve × drive estimate represents the PSQI × reserve × drive interaction. All models were adjusted for age, sex, and resting HR. PSQI and the HRV predictors were standardized; symptom outcomes remained on their original 0–10 VAS scales. All 15 single-index confidence intervals included zero. The joint three-way interaction excluded zero only for phantom limb pain.

### Reserve and drive jointly moderate the sleep–pain relationship

Entered jointly, the three-way interaction of PSQI with autonomic reserve and autonomic drive was significant for PLP (b = −0.51, 95%CI −0.95 to −0.062; F(1,110) = 5.10, p-value = 0.026). The corresponding term was non-significant for RLP (b = −0.44, 95%CI −0.98 to 0.098; F(1,110) = 2.62, p-value = 0.108), in the same direction, and clearly null for PLS (b = 0.181, 95%CI −0.42 to 0.79; F(1,110) = 0.34, p-value = 0.556). The moderation is specific to painful post-amputation symptoms and does not extend to non-painful phantom perception arising from the same deafferented territory.

The negative three-way coefficient indicates that the association between sleep quality and PLP changed differently across autonomic drive depending on reserve. Displayed by reserve stratum, the model-implied slope for PLP declines with increasing drive among participants with preserved reserve and rises among those with reduced reserve (Figure 4). Reserve was retained as a continuous variable in the inferential model, and the median-defined strata are presented only to illustrate the interaction. The conditional PSQI × drive interaction did not reach statistical significance at either representative reserve value (preserved reserve, p-value = 0.054; reduced reserve, p-value = 0.138). However, the three-way interaction tests whether the PSQI × drive interaction changes across reserve values, not whether either conditional interaction separately differs from zero.

**Figure 4.**
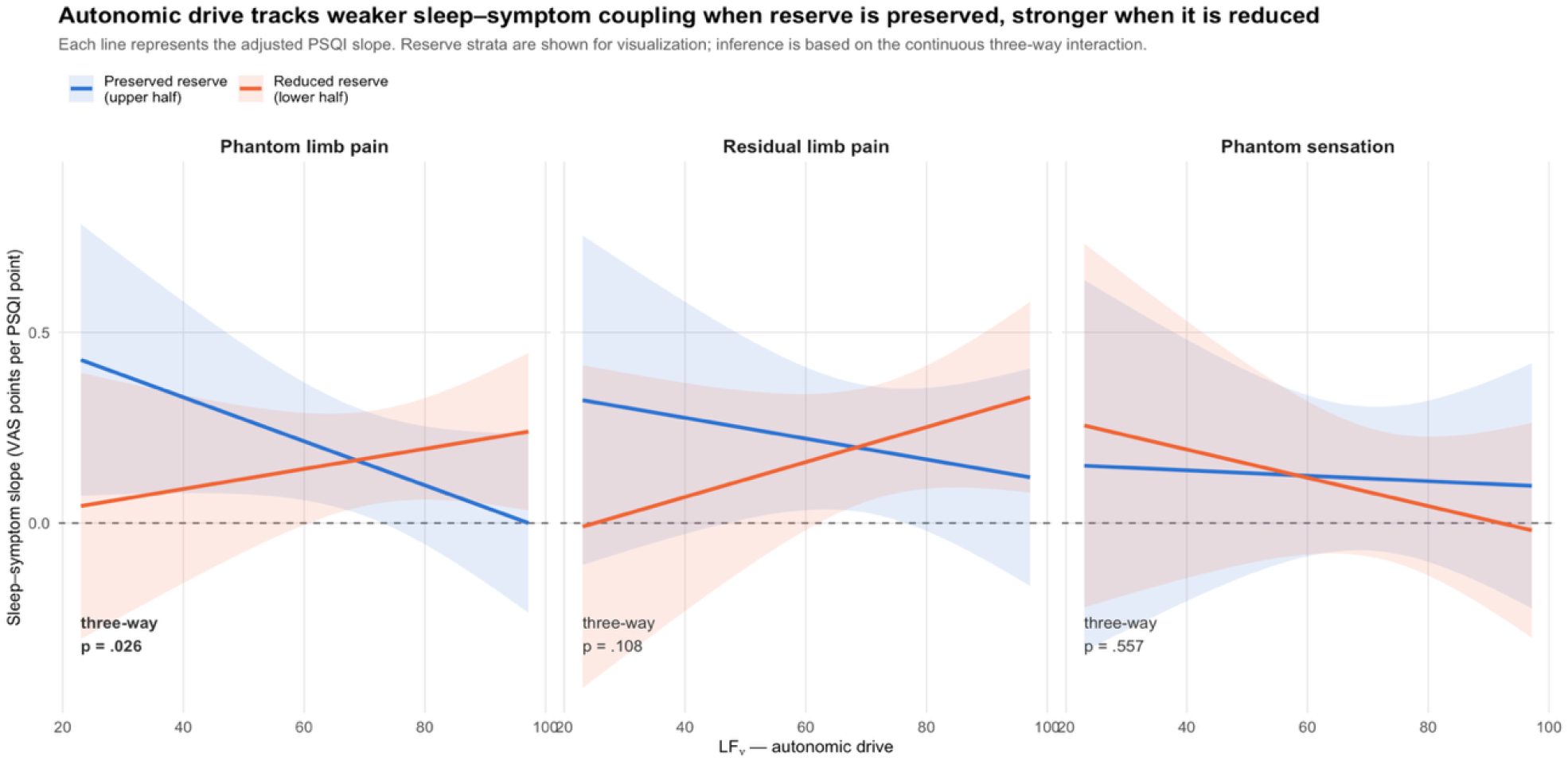
Conditional associations between sleep quality and post-amputation symptoms across autonomic drive and reserve. Figure 4: Lines represent the estimated PSQI slope across LF_nu_ at the median reserve value within the lower and upper halves of the reserve distribution; shaded bands represent 95% CIs. Models were adjusted for age, sex, and resting HR. Reserve was analyzed continuously in all inferential models. The median-defined strata are presente only to visualize the PSQI × reserve × drive interaction. A positive slope indicates that poorer sleep was associated with greater symptom intensity.

### Decomposition: reserve is informative only when drive is elevated

The interaction can be re-expressed as the effect of reserve on the sleep–pain slope at each level of drive (Table 2). Among participants with higher autonomic drive, the adjusted PSQI– PLP slope was essentially flat among participants with preserved reserve (b = −0.03) but positive among those with reduced reserve (b = 0.30). The difference between reserve groups was statistically significant (difference = −0.34, 95% CI −0.64 to −0.03, p-value = 0.03). Among participants with lower drive, reserve made essentially no difference (difference = 0.06, 95% CI −0.24 to 0.37, p-value = 0.65). Autonomic reserve is thus informative about the sleep–pain relationship when compensatory drive is elevated and uninformative when it is not.

**Table 2.**
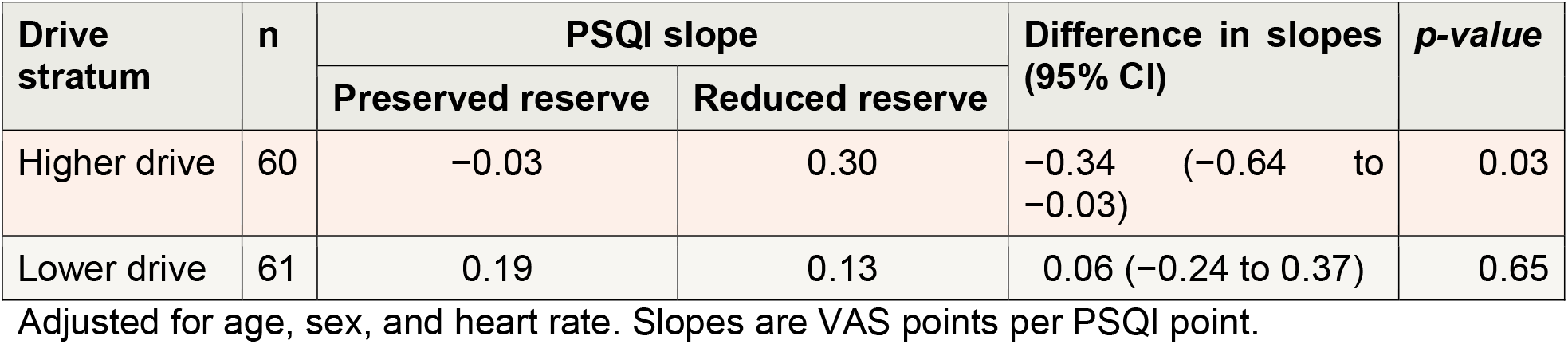
Effect of reserve on the sleep–phantom limb pain slope, at each level of autonomic drive.

| Drive stratum | n | PSQI slope |  | Difference in slopes (95% CI) | p-value |
| --- | --- | --- | --- | --- | --- |
|  |  | Preserved reserve | Reduced reserve |  |  |
| Higher drive | 60 | −0.03 | 0.30 | −0.34 (−0.64 to −0.03) | 0.03 |
| Lower drive | 61 | 0.19 | 0.13 | 0.06 (−0.24 to 0.37) | 0.65 |

Adjusted for age, sex, and heart rate. Slopes are VAS points per PSQI point.

This decomposition is presented as an interpretation of the three-way interaction. The difference between preserved and reduced reserve under higher autonomic drive depended on how reserve was constructed: across the nine reserve definitions examined, the corresponding p-values ranged from 0.036 to 0.147 and reached conventional statistical significance only for the primary reserve index (Supplementary Table 2). In contrast, the continuous three-way interaction, PSQI × reserve × drive coefficient, remained negative and similar in magnitude across all nine definitions, with p-values ranging from 0.021 to 0.057. Inference rests on this interaction, not on single median-stratified comparison.

### Descriptive subgroups

Dichotomizing reserve and drive dimensions at their medians yielded four subgroups whose adjusted sleep–post-amputation symptoms slopes follow the anticipated framework order (Table 3, Figure 5). For phantom limb pain, the adjusted sleep–pain slope was shallowest among participants with preserved reserve and higher drive (b = 0.050) and steepest among those with reduced reserve and higher drive (b = 0.286). The latter slope differed from zero within its subgroup (p-value = 0.004). However, statistical significance within one subgroup does not establish that its slope differs from those of the other subgroups; formal evidence for moderation rests on the continuous PSQI × reserve × drive interaction reported above.

**Table 3.**
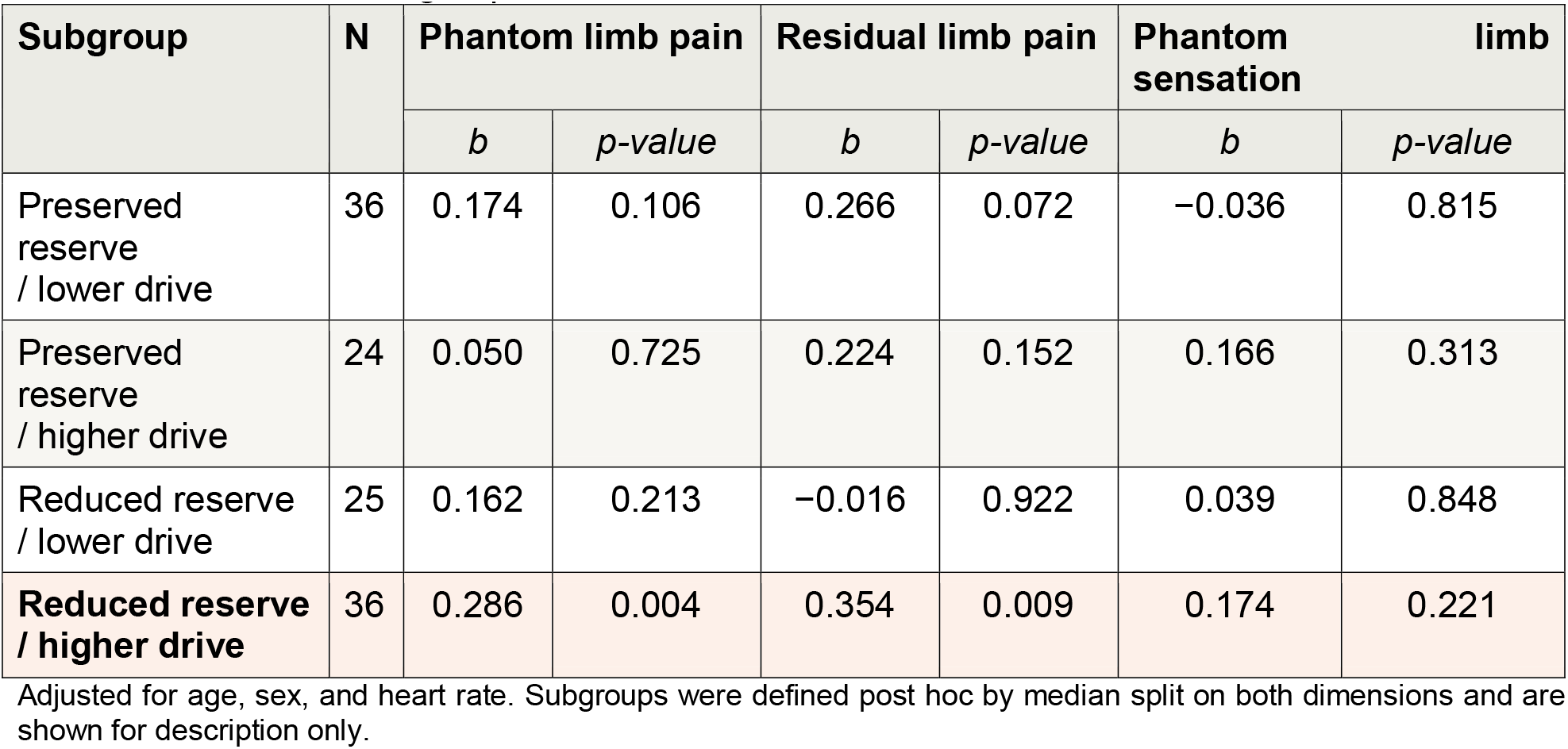
Adjusted sleep–post-amputation symptom slopes (VAS points per PSQI point) within each reserve × drive subgroup.

**Figure 5.**
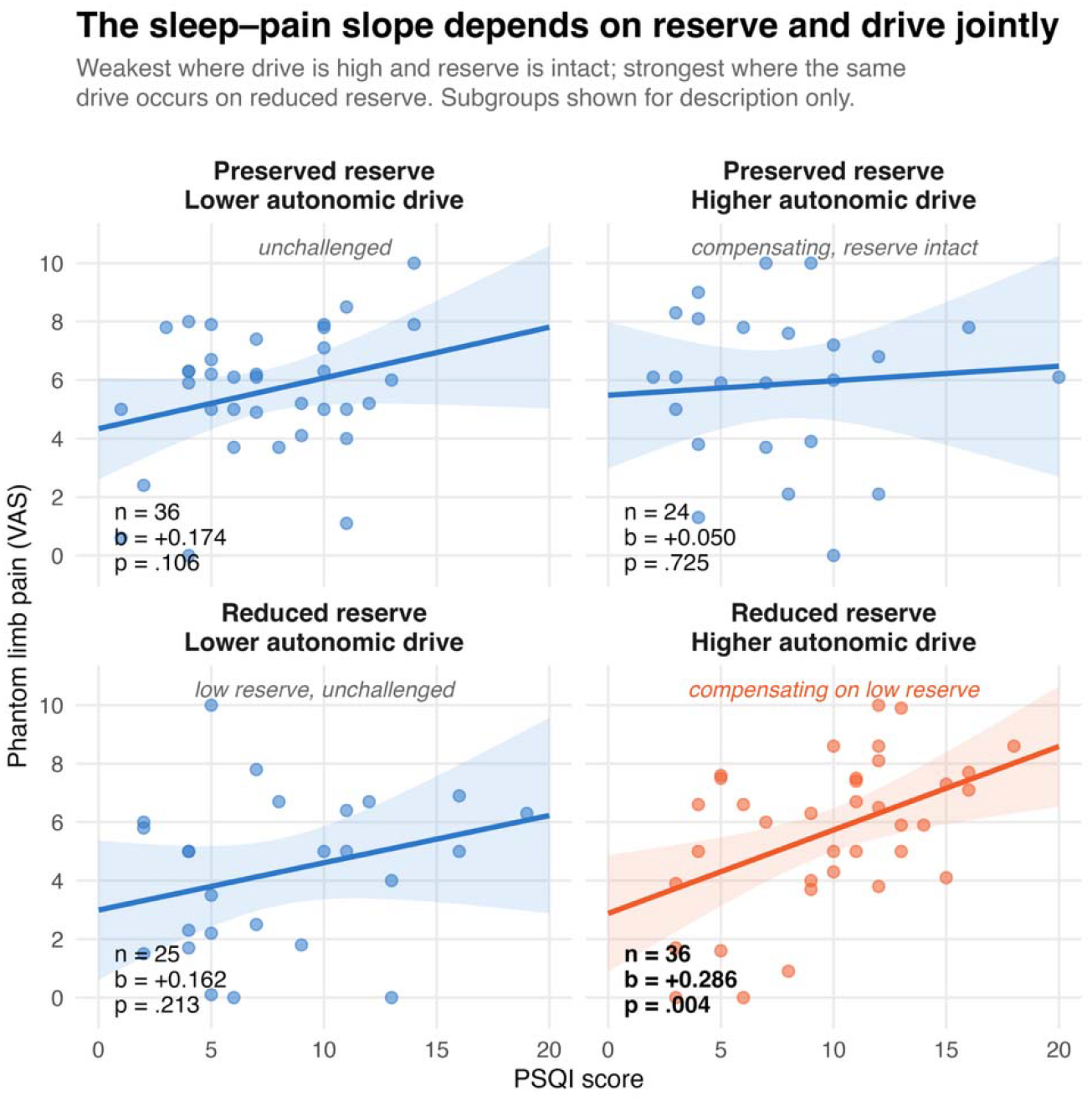
Sleep and phantom limb pain within each reserve × drive subgroup. Figure 5: Points represent observed values; lines and shaded bands represent fitted regression estimates and 95% CIs from models adjusted for age, sex, and resting heart rate. Higher PSQI scores indicate poorer sleep. Autonomic reserve and drive were dichotomized at their respective sample medians for descriptive visualization; drive was indexed by LFnu. The coefficient b represents the adjusted change in phantom limb pain VAS score per one-point increase in PSQI, and p is the nominal two-sided p-value for the within-subgroup slope.

### Sensitivity analyses

The primary three-way interaction remained similar after additional covariate adjustment. With BDI and BAI included, the interaction was b = −0.466, p-value = 0.047, and with current pain medication use and time since amputation additionally included, the interaction was b = −0.475, p-value = 0.045. The interaction was also robust to the construction of the reserve index (across the nine definitions), remaining significant under every alternative definition examined except RMSSD alone (range p-values from 0.021 to 0.057; Supplementary Table 2). This included definitions showing negligible empirical correlation with drive, such as SDNN plus TP (r = 0.017; interaction p-value = 0.026) and SDNN alone (r = −.023; interaction p-value = 0.033), suggesting that the interaction is not an artifact of that overlap. Substituting ln(LF/HF) for LFnu as the drive index produced a similar result (b = −0.488, p-value = 0.029). A percentile bootstrap with 3,000 resamples produced a 95% CI of −1.057 to −0.050, with 98.5% of bootstrap estimates below zero. Taken together, the direction and magnitude of the interaction were stable across covariate sets, nine reserve definitions, an alternative drive metric and bootstrap resampling, and the effect was specific to the painful phantom outcome.

## Discussion

To our knowledge, this is the first study to examine the sleep–phantom limb pain relationship within a multidimensional autonomic framework that jointly considers regulatory capacity and concurrent autonomic state. Rather than relying on individual HRV measures, we characterized autonomic reserve using a composite of global HRV magnitude and autonomic drive using normalized low-frequency power. This approach tested whether the association between sleep disturbance and pain depends on the physiological context in which autonomic regulation occurs. Several findings emerged. Poorer sleep quality was associated with greater PLP and RLP, but not with non-painful PLS. For RLP, the association appeared to be driven by the presence of pain rather than by its intensity. No individual HRV index moderated these associations. In contrast, the joint interaction of sleep quality with autonomic reserve and autonomic drive was significant for PLP, with the steepest sleep–pain slope in participants who were mounting a high drive from a reduced reserve, and a flat slope in those mounting the same drive from a preserved reserve. This pattern was stable across covariate adjustment, nine definitions of reserve, an alternative drive metric, and bootstrap resampling, and it was specific to painful phantom experience.

The central result is that the autonomic correlate of vulnerability to sleep-related pain is conditional, not linear. Reduced reserve was not by itself associated with a stronger sleep–pain relationship, and neither was elevated drive; reserve became informative only when drive was elevated (Table 2), and at low drive it made no difference. This is precisely the pattern that a compensation model predicts and that a single-marker model cannot produce. The logic is that of glucose regulation: an insulin concentration says nothing about β-cell health until it is read against the glycemia it is sustaining, and the informative quantity is therefore insulin relative to glucose, not either alone^25,26^. Cognitive reserve carries the same structure, in that the same pathology produces different clinical expression depending on the capacity available to compensate for it^27^, as does allostatic load, in which the cost of a compensatory response depends on whether it can be sustained within the capacity of the system^28^. Read in these terms, our participants with high drive and preserved reserve were compensating effectively, and disturbed sleep did not translate into pain; those with high drive and reduced reserve were compensating at diminishing efficiency, and the same degree of sleep disturbance did. Low drive with low reserve was not associated with a steep slope because a capacity that is not being drawn on is not being tested.

These findings converge with what we have observed at the cortical level. In fibromyalgia and knee osteoarthritis, the analgesic efficiency of motor cortex inhibition depends on the burden of comorbid stressors a patient carries: the same inhibitory signature is associated with less pain when neurocompensatory capacity is preserved and with more pain when it is saturated, a relationship we formalized as a resistance index analogous to HOMA-IR and that recalibrates with successful rehabilitation^24^. The present study extends this logic from the cortical to the autonomic axis and from a between-patient burden phenotype to a within-system measure of capacity. In both cases, the decisive quantity is not the size of the compensatory response but the response relative to the reserve supporting it, and in both cases the failure mode is the same: a system that keeps responding, but at diminishing efficiency. Reduced HRV reserve under high drive is, in this sense, the autonomic signature of compensatory resistance, in the way that hyperinsulinemia with hyperglycemia is the metabolic one. That HRV also predicts β-cell compensatory capacity in vivo^22^ suggests that these are not parallel analogies but expressions of a shared regulatory reserve, an idea that a combined cortical and autonomic assessment in the same patients could now test directly.

This framework also offers an explanation for a long-standing inconsistency in the literature. Meta-analyses generally report lower global and vagally mediated HRV in chronic pain, yet effect sizes vary widely across conditions, HRV correlates with pain intensity inconsistently^31,32^, and studies in people with limb amputation likewise suggest altered autonomic regulation without a simple correspondence between any HRV measure and PLP severity^18,19^. If autonomic variability matters as a constraint on the response to additional perturbation rather than as a direct correlate of pain, then no single index should be expected to track pain linearly, any more than a fasting insulin concentration should be expected to track glycemia. The strongest signal should arise from an interaction, which is what we observed. Reduced reserve does not generate pain by itself; it narrows the margin within which the organism can absorb competing regulatory demands, such as disturbed sleep, without amplifying symptom expression.

What the two axes measure deserves precision. The reserve index integrates SDNN, RMSSD, TP, and HF-power, which were highly collinear in this sample, so the composite captures a single global-variability dimension, not several independent autonomic processes. This is consistent with the substantial overlap among conventional HRV measures and with prior composite approaches^23,30^. It maps onto the distinction drawn by Berntson and colleagues between autonomic balance, which describes the current state, and regulatory capacity, which describes what the system can sustain^21^. Reserve, on this reading, is capacity. Drive, indexed by LFnu, describes the relatively low-frequency organization of the spectrum and not sympathetic activity: normalized spectral measures are shaped by baroreflex dynamics, vagal modulation, respiration and the arithmetic of normalization^20,33^. The data therefore do not show that sympathetic activation is harmful only when reserve is low. They show that the same spectral organization carries a different meaning for the sleep–pain relationship depending on the magnitude of variability available to support it. That the interaction survived reserve definitions excluding HF power, including those with negligible correlation with drive, makes mathematical overlap between the axes an unlikely explanation.

The specificity of the interaction to PLP supports interpreting reserve in relation to pain regulation rather than phantom representation in general. PLP and PLS arise from the same deafferentation but are not equivalent phenomena. Contemporary models place PLP at the intersection of peripheral, spinal, cortical and affective-motivational processes, whereas a phantom percept can persist without becoming painful^34,35^. Neither the direct sleep association nor the reserve × drive moderation was present for PLS, so reduced reserve did not simply track the persistence or vividness of the phantom. The findings are more compatible with a role in the processes that determine whether ongoing post-amputation activity is expressed as pain, through nociceptive gain, salience, arousal, descending modulation, or the capacity to recover from repeated perturbation. The RLP result, in which sleep distinguished those with any pain from those with none but not intensity among the affected, is consistent with the same threshold logic.

The mechanism is biologically plausible because autonomic control and pain regulation share their central substrate. The insula, anterior cingulate cortex, amygdala, hypothalamus, periaqueductal gray, and brainstem nuclei contribute to both^36,37^; cardiac autonomic dynamics are tightly coupled to sleep architecture and cortical state^38^; and nociceptive input during sleep evokes measurable autonomic responses even without behavioral awakening^16^. Sleep disturbance can thus be understood as a recurrent demand on a system that is already engaged in pain control. Experimental sleep loss increases pain sensitivity and alters endogenous modulation, although effects on conditioned pain modulation are not uniform^9,39^.

Whether that demand is absorbed or expressed as pain would then depend on the reserve available to meet it, which is what the interaction shows.

Some limitations should be weighed. The design is cross-sectional, so the direction of the sleep–pain relationship and the causal role of reserve cannot be established; the prospective phase of the parent trial will allow us to test whether reserve changes with treatment and whether such change tracks clinical response, as cortical compensatory efficiency does^24^. Inference rests on a single prespecified interaction test at p-value = 0.026 in 121 participants, and although the effect was robust to every sensitivity analysis we ran, a three-way interaction of this magnitude requires replication in an independent cohort before the framework can be considered established. We have reported nominal p-values and made the analytical decisions explicit so that such replication can be exact. HRV was derived from a single five-minute resting recording, which captures capacity at one moment rather than its dynamics. The reserve composite was constructed for this analysis and, like any formative index, will need external validation; the drive index is an interpretation of spectral organization and not a measure of sympathetic outflow. Median-defined subgroups were used for description only, and the within-subgroup coefficients should not be read as separate tests. Finally, the sample comprised trial participants with chronic PLP of at least moderate intensity, and generalization to individuals with mild or absent PLP is untested.

In summary, the relationship between autonomic function and PLP appears to be conditional rather than linear. Reduced autonomic reserve was not simply associated with greater pain; it became informative under elevated drive, where preserved reserve buffered and reduced reserve amplified the translation of sleep disturbance into pain. This reframes HRV in post-amputation pain from a search for a single biomarker to a model of regulatory capacity relative to demand, the same model that explains why cortical inhibition loses its analgesic efficiency under allostatic load. Because both reserve and drive are modifiable through sleep restoration, aerobic exercise, vagal stimulation, and rehabilitation, the framework points to treatable targets rather than fixed traits, and to the possibility that restoring reserve, rather than suppressing pain directly, is what allows a patient to sleep badly without paying for it in pain.

## Supporting information

Strobe Checklist

## Author contributions

Conception and design: FF and NA. Data access: FF, NA and MH. Data cleaning and pre-processing: NA and MH. Data analysis: FF and NA. Interpretation of the results: All authors. Writing the first draft of the manuscript: All authors. Reviewed and approved the final version of the manuscript: All authors.

## Funding

This study was supported by an NIH R01 grant (5R01HD082302-09). The funder had no role in study design, data collection, analysis, interpretation, or the decision to submit the manuscript for publication.

## Data availability

The de-identified participant-level data supporting the findings of this study are available from the corresponding author on reasonable request, subject to approval by the Mass General Brigham Institutional Review Board and a data-use agreement, because they derive from an ongoing clinical trial.

## Competing interests

The authors declare no competing interests.

## Notes

### Competing Interest Statement

The authors have declared no competing interest.

### Clinical Trial

NCT05188183

### Author Declarations

The parent trial was approved by the Mass General Brigham Institutional Review Board (protocol 2021P003323) and is registered at ClinicalTrials.gov (NCT 05188183).

