## Supplementary Material for "Compensatory drive and sleep-related phantom limb pain: the moderating role of autonomic reserve"

**Supplementary Table 1. Construction diagnostics for the primary reserve index (n=121).**

| **Diagnostic** | **Estimate** |
| --- | --- |
| **Cronbach’s alpha** | 0.98 |
| **Pairwise component correlations, range** | 0.87 – 0.99 |
| **Variance explained by the first principal component** | 94.3% |
| **Correlation between reserve and LFnu** | −0.17 |

**Supplementary Table 2. Primary interaction and higher-drive simple effect under alternative reserve definitions for phantom limb pain.**

| **Reserve definition** | **r with LFnu** | **Three-way b** | **95% CI** | **p-value** | **Higher-LFnu simple effect p-value** |
| --- | --- | --- | --- | --- | --- |
| **SDNN + RMSSD + TP + HF**  **(primary)** | −0.17 | −0.51 | (−0.96, −0.06) | 0.02 | 0.036 |
| SDNN + RMSSD + TP | −0.11 | −0.49 | (−0.94, −0.05) | 0.03 | 0.087 |
| SDNN + RMSSD  (time domains) | −0.19 | −0.47 | (−0.91, −0.03) | 0.04 | 0.067 |
| TP + HF  (frequency domains) | −0.14 | −0.54 | (−0.99, −0.09) | 0.02 | 0.143 |
| SDNN + TP | +0.01 | −0.51 | (−0.96, −0.06) | 0.03 | 0.101 |
| SDNN alone | −0.02 | −0.48 | (−0.93, −0.04) | 0.03 | 0.101 |
| TP alone | +0.06 | −0.54 | (−0.99, −0.08) | 0.02 | 0.129 |
| HF power alone | −0.33 | −0.50 | (−0.94, −0.07) | 0.02 | 0.147 |
| RMSSD alone | −0.35 | −0.43 | (−0.89, 0.01) | 0.05 | 0.112 |

The primary reserve index comprised the mean of standardized natural-log-transformed SDNN, RMSSD, total power, and HF power. All regression models were adjusted for age, sex, and resting heart rate. The three-way interaction is stable across definitions; the simple effect at higher drive is not. LFnu = low-frequency power in normalized units; CI = confidence interval; HF = high frequency; RMSSD = root mean square of successive RR-interval differences; SDNN = standard deviation of normal-to-normal RR intervals; TP = total power.

**Supplementary Figure 1.** Construction of the autonomic reserve index. The four components are near-collinear and behave as a single factor (left). The resulting index is weakly associated with the drive axis (centre), because high-frequency power contributes to both (right).

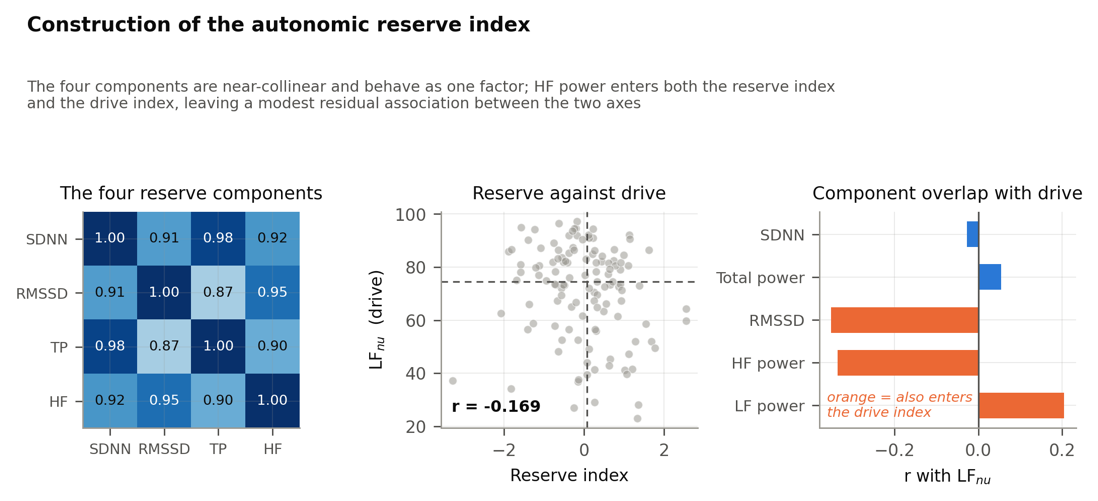

**Supplementary Table 3. Baseline characteristics of the included participants.**

| **Variable** | **Total** |
| --- | --- |
| N | 122 |
| Age, years | 57.55 ± 15.43 |
| Sex, male | 80 (65.6) |
| Amputation-adjusted BMI, kg/m² | 32.22 ± 9.03 |
| Race, white | 100 (82) |
| Ethnicity, not Hispanic or Latino | 111 (91) |
| Time since amputation, years | 4.00 (2.00-7.00) |
| Pain before the amputation, yes | 70 (57.4) |
| Current pain medication, yes | 88 (72.1) |
| PSQI score | 8.28 ± 4.24 |
| PLP intensity (VAS) | 5.43 ± 2.54 |
| RLP intensity (VAS) | 3.09 ± 3.11 |
| PLS intensity (VAS) | 4.68 ± 3.38 |
| Depression symptoms (BDI) | 10.63 ± 6.41 |
| Anxiety (BAI) | 11.44 ± 8.59 |
| Heart rate (bpm) | 76.50 (67.25-85.00) |
| Mean RR interval (ms) | 784.50 (702.50-894.50) |
| SDNN (ms) | 18.40 (11.03-31.00) |
| RMSSD (ms) | 14.95 (8.72-24.23) |
| pNN50 (%) | 0.34 (0.00-5.47) |
| LF power (ms²) | 203.00 (60.25-514.00) |
| HF power (ms²) | 66.50 (20.25-200.75) |
| LF power (nu) | 74.02 (58.59-84.06) |
| HF power (nu) | 25.48 (15.89-41.37) |
| Total power (ms²) | 313.50 (107.00-812.50) |

Data are presented in *mean ± SD, median (IQR), or absolute and relative (%) frequencies.* Percentages were calculated using the number of non-missing observations for each variable. BMI, body mass index; HF, high frequency; HRV, heart rate variability; LF, low frequency; PSQI, Pittsburgh Sleep Quality Index; RMSSD, root mean square of successive differences; SDNN, standard deviation of normal-to-normal intervals.

**Supplementary Table 4. Constituent Heart Rate Variability Measures by Median-Defined Autonomic Reserve Group.**

| **Variable** | **Total** | **Lower Median** | **Higher Median** |
| --- | --- | --- | --- |
| SDNN (ms) | 18.40 (11.03-31.00) | 10.9 (8.1-14.4) | 31.2 (23.4-40.3) |
| RMSSD (ms) | 14.95 (8.72-24.23) | 8.8 (5.1-11.1) | 24.3 (18.9-38.6) |
| HF power (ms²) | 66.50 (20.25-200.75) | 20.0 (9.0-32.0) | 202.0 (118.0-316.0) |
| Total power (ms²) | 313.50 (107.00-812.50) | 105.0 (58.0-173.0) | 814.0 (488.0-1431.0) |
